# Association of Vaccine Uptake and COVID-19 Infections among nursing home staff and residents in Missouri, United States

**DOI:** 10.1101/2021.09.16.21263714

**Authors:** Stephen Scroggins, Matthew Ellis, Enbal Shacham

**Affiliations:** Saint Louis University, College for Public Health and Social Justice & The Geospatial Institute, St. Louis, MO, USA; Washington University in St. Louis, School of Medicine, Department of Psychiatry, St. Louis, MO, USA

## Abstract

Nursing homes (NH) continue to struggle with COVID-19 morbidity and mortality with older adult residents at greater risk of infection due to proximity to other residents, advanced aging-related chronic illnesses, and contact with staff. While many states have prioritized COVID-19 vaccinations among older adults, vaccinations among NH staff vary. The purpose of this study was to quantify the relationship between nursing home staff vaccination uptake and COVID-19 infections among residents. A zero-inflated Poisson regression model was constructed to predict the weekly number of COVID-19 cases among Missouri nursing home residents using data from the Centers for Medicaid and Medicare Services. A total of 1,124 COVID-19 infections were reported among 504 NH residents between January 1, 2021 and August 22, 2021. After adjusting for number of total residents, resident vaccine rate, staff quality rating, and respective county COVID-19 rate, for every percent increase in nursing home staff vaccine rate the risk of COVID-19 infections significantly decreased by 13% (IRR 0.87, 95% CI 0.81, 0.93). This study identified that NH staff, likely due to greater mobility, are important to prioritize in vaccination efforts to protect themselves and residents of their facilities from COVID-19 infections. Further, the CMS staff ratings were significant predictors of infection as well, which highlight the structural challenges that exist within and outside the context of a highly infectious and deadly pandemic. These results also provide insights to optimizing vaccination roll-out to best protect our communities’ most vulnerable residents.

## Introduction

In the United States, COVID-19 infections continue to spread, with outcomes disproportionately affecting certain populations.^1^ In particular, older individuals (i.e., those over the age of 60) are at increased risk of morbidity and mortality associated with COVID-19 due to widely prevalent pre-existing conditions such as cardiovascular disease and hypertension.^2-4^ Within this population of older adults, those living in communal spaces such as nursing or retirement communities are at even greater risk of infection due to close proximity to others, more advanced aging-related or medical conditions, and engagement with staff that may support multiple clients and facilities.^4-5^ In addition, mobility between residents and facilities provides opportunities for staff to act as potential vectors of COVID-19. Throughout the pandemic, reports have noted COVID-19 outbreaks occurring at nursing home (NH) facilities throughout the U.S..^6^

As a result, the Centers for Disease Control and Prevention (CDC) has established NH-specific COVID-19 infection control strategies (e.g., routine testing, personal protective equipment, respiratory protection programs).^7^ Additionally, the CDC has prioritized healthcare personnel and older individuals as first line recipients of the COVID-19 vaccine. However, NH healthcare staff may not have been initially perceived in the same manner as those working in hospital settings or those who are credentialed, professional staff (i.e., nurses, physician assistants).

In addition to control strategies, vaccine uptake has been shown to reduce the spread of COVID-19 in the general population.^8^ However, despite being a high-risk population, there is little data on vaccine uptake and outcomes among NH residents, and the role staff vaccine uptake may play on continued COVID-19 risk among this population. Importantly, how vaccine uptake may have differed, and whether vaccine uptake is more effective in mitigating fatal outcomes when focused on staff, residents, or both. These analyses are crucial for informing organizational and public policy applicable to COVID-19 within NHs amidst an ongoing pandemic.

## Methods

### Study Design and Sample

Data for this study were extracted from two publicly available sources. Centers for Medicaid and Medicare Services (CMS) houses a longitudinal dataset contains characteristics of NHs, weekly reported observations of residential COVID-19 cases, and COVID-19 vaccine coverage of healthcare staff and residents from facilities that utilize reimbursement for public insurance sources.^8^ Second, as COVID-19 infection rates in NHs may be associated with rates in the surrounding county, data from Missouri Department of Health and Senior Services (MDHSS) were extracted and aggregated to calculate the average weekly COVID-19 rate for each Missouri county.^9^

Weekly observations for this study were limited to NHs located within the state of Missouri and occurring between May 24, 2021 and August 29, 2021, a period when vaccine distribution across the state and country was more widely prevalent than earlier in the year.

### Measures

The weekly confirmed COVID-19 case count among residents per NH was the primary study outcome. New weekly cases of COVID-19 infections were identified among residents of NHs, confirmed with one of several diagnostic tests. Observations with missing values were excluded from analysis.

The primary predictors of interest for this study were: 1) the proportion of healthcare staff (e.g., exclusive of maintenance or auxiliary staff) per NH who had received a partial/complete COVID-19 vaccination at the time each observation was made; and 2) the proportion of residents per NH who had received a partial/completed COVID-19 vaccination at time of each observation. These measures, calculated by CMS, utilize the total number of staff and residents per NH, reported each week, as the denominator. For observations with missing values, the respective NH average was used.

Additional a priori predictor variables included were: 1) total number of residents; 2) weekly average COVID-19 rate of the county where the NH resides; and 3) most recent CMS staff rating was used as covariates. To the latter point, CMS assigns NHs a composite rating, 1 being weakest and 5 being strongest, of staff adequacy based on met needs of residents and average number of daily hours of care provided to each resident by staff.

### Statistical Analysis

Descriptive statistics of weekly COVID-19 cases among NH residents along with a priori variables were assessed.A zero-inflated Poisson (ZIP) regression model was constructed to predict (1) if COVID-19 was reported, and if so, (2) the number of cases reported. ZIP regression was used due to the count nature of the outcome and due to the pre-determined overdispersion of observations when zero COVID-19 cases were reported during the study period.

## Results

A total of 1124 cases of COVID-19 were reported among residents among 504 Missouri NHs during the study period. Table 1 details NH characteristics.

**Table 1.** Missouri NH Descriptive Statistics and Correlation to Weekly COVID-19 Case among Residents (N=504)

|  |  |  |
| --- | --- | --- |
| Characteristic | Weekly Average (SD) | 25 and 75 Interquartile |
| Proportion of NH staff partially or completely COVID-19 vaccinated | 48.2 (19.7) | 34.2, 62.2 |
| Proportion of residents partially or completely COVID-19 vaccinated | 84.3 (15.1) | 79.4, 94.0 |
| Average number of weekly residents in NH | 66.2 (34.9) | 44, 81 |
| County COVID-19 rate per 1,000 residents | 25.4 (21.8) | 6.3, 40.6 |
| Staff rating | 2.8 (1.2) | 2, 4 |

Each week, there was an average of 48.2% (SD=19.7) of NH staff and 84.3% of residents (SD=15.1) were partially or completely vaccinated across Missouri. NHs had a weekly average of 66 residents staying at the facility (SD= 34.9), with a range of 2 to 100. Among the 115 counties in Missouri, the average COVID-19 rate was 25.4 per 1,000 (SD=21.8) and ranged from 0 to 127.6 per 1,000 throughout the study period. On a scale of 1 to 5, the average staff rating of NHs was 2.8 (SD=1.2).

Table 2 details the model exponentiated coefficients of the zero inflated Poisson regression. First, the Bernoulli process indicates that as the number of NH residents increases, the odds of COVID-19 infections being reported significantly increased by 1% (AOR 0.99, 95% CI 0.99, 0.99) and as the county rate of COVID-19 infections increased the odds of COVID-19 being reported among NH residents significantly increased by 3% (AOR 0.97, 95% CI 0.97, 0.98).

**Table 2.** Zero Inflated Poisson Regression Model Predicting Weekly COVID-19 Cases Among Missouri NHs Residents (n=504)

|  |  |  |  |
| --- | --- | --- | --- |
| Part 1: Zero-logistic (Bernoulli Logit Process) |  |  |  |
|  | Adjusted Odds Ratio | 95% CI | P-Value |
| Number of residents | 0.99 | 0.99, 0.99 | 0.007 |
| County COVID-19 rate | 0.97 | 0.97, 0.98 | <0.001 |
| Staff rating | 0.94 | 0.85, 1.03 | 0.180 |
| Percentage of NH staff any COVID-19 vaccination | 1.00 | 0.99, 1.01 | 0.066 |
| Percentage of residents any COVID-19 vaccination | 0.99 | 0.98, 1.00 | 0.137 |
| Part 2: Count (Poisson Log Process) |  |  |  |
|  | Incidence Risk Ratio | 95% CI | P-Value |
| Number of residents | 1.00 | 0.99, 1.00 | 0.353 |
| County COVID-19 rate | 1.00 | 1.00, 1.01 | 0.008 |
| Staff rating | 0.87 | 0.81, 0.93 | <0.001 |
| Percentage of NH staff any COVID-19 vaccination | 0.99 | 0.98, 0.99 | 0.008 |
| Percentage of residents any COVID-19 vaccination | 0.99 | 0.99, 1.01 | 0.153 |

The count process of the regression model indicates significant associations in part 2 of the model to understand predictors of when NHs experienced COVID-19 infections during the study period. As county COVID-19 rate increased, the risk of COVID-19 incidence among NH residents significantly increased (IRR 1.0, 95% CI 1.00, 1.01). As NH staff rating increased, the risk of COVID-19 infection among residents decreased by 13% (IRR 0.87, 95% 0.81, 0.93). Finally, as the percent of vaccinated staff increased, the risk of COVID-19 infections among residents significantly decreased (IRR 0.99, 95% CI 0.98, 0.99). Percent of residents vaccinated against COVID-19 was not significantly associated with whether COVID-19 cases were reported, nor the number of COVID-19 infections reported.

## Discussion

The purpose of this study was to examine whether COVID-19 vaccinations protected NH residents by analyzing staff and resident rates of vaccination. These results highlight the importance of prioritizing vaccine uptake among NH staff as a method of protecting resident populations, in addition to themselves. Better understanding of barriers to staff vaccination and hesitancy is urgent; further, increasing communication efforts to increase uptake is necessary. Lastly, developing and implementing comprehensive infection control protocols during a pandemic for NHs has been challenging, and these findings should be incorporated into future policy planning, particularly prioritizations for booster vaccination shots.

Importantly, this study identified that when NH staff had a lower vaccination rate, there were higher COVID-19 infection rates in nursing homes, which may be explained by community mobility. NH staff members move throughout their days, not only within single or multi-site organizational spaces as a matter of employment, but also throughout one’s non-employment-related life. Thus, there are increased opportunities for COVID-19 infection and subsequent transmission compared to less mobile and more isolated residential populations of NHs. However, despite the identification of healthcare personnel within NH communities as “essential” in terms of continued employment, and the limitations of other persons such as family and friends from engaging with NH residents, this role was not prioritized as a possible vector of infection transmission among vulnerable populations, older adults in organized, community settings. While such employees were encouraged to become vaccinated, there were not particular, systematic efforts to enhance support for workers that place themselves at COVID-19 risk. Furthermore, the Biden administration has recently implemented a requirement where that all staff at NHs will receive a COVID-19 vaccine to receive continued Medicare funding, this signals the support of these type of data and results.

Interestingly, this study also identified that NH staff ratings were important predictors of COVID-19 infections; higher ratings suggested lower number of reported COVID-19 infections among NH residents. While this measure may serve as a proxy for detailed care of NH residents, including adhering to infection control protocols, it may also be true that appropriate staffing resulted in fewer resident contacts per staff member and subsequently, less opportunity for COVID-19 transmission. These data highlight structural challenges that may exist outside of the context of a pandemic and provide validation that this measure can and should be used as a tool for intervention to improve health outcomes of the residents in other domains.

In conclusion, these findings highlight the need to prioritize vaccinations in community members who are more mobile throughout their daily lives, in this case NH healthcare staff. These findings inform policy and can justify the need to prioritize front line workers that provide care for vulnerable populations. Future studies should incorporate community mobility as an integral component of COVID-19 risk assessment.

## Data Availability

All data used within this study is publically available and has been thoroughly cited.

## Acknowledgement

This study was partially funded by the Sinquefield Center for Applied Economic Research.

